# Age-related differences in colon and rectal cancer survival: An analysis of United States SEER-18 data

**DOI:** 10.1101/2022.11.29.22282871

**Authors:** Sophie Pilleron, Diana Withrow, Brian D Nicholson, Eva JA Morris

## Abstract

Age-related differences in colon and rectal cancer survival have been observed, even after accounting for differences in background mortality. To determine to what extent stage, tumour site, or histology could contribute to these differences, we estimated 1-year relative survival (RS) age stratified by these factors. Colon and rectal cancer cases diagnosed between 2012 and 2016 and followed up until 2017 were retrieved from 18 United States Surveillance Epidemiology and End Results cancer registries. For colon cancer, 1-year RS ranged from 87.8% [95% Confidence Interval: 87.5-88.2] in the 50–64-year-old age group to 62.3% [61.3-63.3] in the 85–99-year-old age group and for rectal cancer ranged from 92.3% [91.8-82.7] to 65.0% [62.3-67.5]. With respect to stage, absolute differences in RS between 50–64-year-old and 75–84-year-old in RS increased with increasing stage (from 6 [5-7] %-points in localized disease to 27 [25-29] %-points in distant disease) and were the highest for cancers of unknown stage (>28%-points). With respect to topography, age-related differences in survival were smallest for those in right-sided colon (8 [7-9] %-points) and largest for tumours of the colon without topography further specified (25 [21-29] %-points). While age-related differences in survival varied by histology and tumour site, the overall age-related differences in survival could not be explained by differences in distribution of these factors by age, consistent with a hypothesis that stage at diagnosis or treatment are more likely drivers. Incorporating data on geriatric conditions such as frailty and comorbidity would support further understanding of the age gap in colon and rectal cancer survival.

## Introduction

In the United States (US), an estimated 150,000 new colon and rectal cancers were diagnosed and over 50,000 deaths occurred in 2021.^1^ Over 30% of colorectal cancers are diagnosed in persons aged 75 and older.^2^ The 5-year relative survival for colon and rectal cancer is estimated to be 64% in the US, similar to that in other high-income countries.^3^ Older adults have poorer cancer survival than middle-aged adults even after accounting for differences in life expectancy. The difference has widened over time, with generally greater improvements in survival noted in younger than older age groups, suggesting older adults have not benefited to the same extent as younger adults from improvements in cancer treatment over recent decades.^3^

Recent studies of age-related differences in colon and rectal cancer survival in high-income countries have shown decreasing survival with increasing age, and larger age-related differences in cancer survival with worsening stage at diagnosis.^3-5^ To date, these studies have not explored whether age-related differences in survival are consistent across histology and/or anatomical tumour site. In this study, we sought to fill this gap, motivated by the hypothesis that further understanding of the epidemiology of survival differences by age could inform interventions. Specifically, using US population-based cancer registry data, we describe age-related differences in relative survival (RS) for colon and rectal cancers by stage at diagnosis, sex, tumour site, and histology.

## Methods

### Data Source

We included all first primary colon and rectal cancers (ICD-10: C18-20) diagnosed between 2012 and 2016 from 18 US Surveillance Epidemiology and End Results (SEER) cancer registries with vital status through December 2017. Cancers were originally coded using ICD-O-3 and grouped according to the SEER ICD-O-3/WHO 2008 definitions.^6^ The 18 SEER registries are located in California (San Francisco/Oakland, San Jose/Monterey, Los Angeles, Greater California), Connecticut, the Detroit metropolitan area, Hawaii, Iowa, New Mexico, Seattle, Utah, Georgia (Atlanta, rural Georgia and Greater Georgia), Alaska (restricted to Alaska Natives), Kentucky, Louisiana, and New Jersey.^7,8^ Together, these population-based registries cover nearly 30% of the US population.^7^

Because early onset colon and rectal cancer has different features and management, we restricted to cases diagnosed in patients aged over 50.^9-11^ We excluded cancers that were registered based on death certificate or autopsy only (1.3%), were non-malignant (3.8%), were missing age (0.08%), or with disagreement between vital status and survival time (0.2%).

### Analysis

We estimated 1-year relative survival (RS) and 1-year RS conditioning on surviving 1 year using the Ederer II method. Relative survival is a metric of net survival, which estimates the probability of survival from cancer in the hypothetical scenario where people cannot die from other causes of death. This metric is useful for comparing survival between groups of people for whom background mortality differs. It is estimated based on the ratio of the observed survival to the expected survival of persons of similar demographics in the general population^12,13^.

Expected survival was estimated from life tables stratified by county, age, year, sex, race/ethnicity and county-level socioeconomic status (SES) index^14^. The SES index is a composite variable based on seven county-level SES attributes collected by the American Community Survey^15^.

RS estimates were stratified by age (50-64, 65-74, 75-84, and 85 to 99), stage (SEER Summary Stage, based on collaborative staging and categorized as localized, regional, distant, and unknown/unstaged^16^), sex, anatomical tumour site (ICD-O-3 topography: right-sided colon: C180-184, left-sided colon: C185-187, C199, colon, NOS: C188-189), and histology (ICD-O-3, adenocarcinoma [8255-8263], mucinous adenocarcinoma [8470, 8480, 8481], signet cell carcinoma [8490]). Patients aged 100+ were excluded since this is the cut-off for expected life tables.^17^

We estimated the absolute differences in RS between the 50-64 years age group and 75-84 years age group. 95% confidence intervals around RS differences were obtained using Monte Carlo simulations assuming the complementary log-log transformed RS values to be normally distributed.^18^ For each difference considered, the confidence limits were obtained by taking the 2.5 and 97.5 percentiles of the empirical RS difference distribution resulting from 100,000 random draws in the complementary log-log transformed RS values distribution.

The 75 to 84 age group was chosen as the comparison group because remaining life expectancy for this age group ranges from 6 years (for men aged 84) to 13 years (for women aged 75) years (https://www.ssa.gov/oact/STATS/table4c6.html) and therefore is an age group for whom improvement in cancer survival may be more achievable and impactful than among the oldest adults.

SEER data are publicly available and ethics approval was not required. Data and cancer survival estimates were retrieved through SEER*Stat software version 8.3.9 (National Cancer Institute). Data was extracted between February and April 2022 by DW, and SP analysed data using R statistical software (version 4.0.2; R Development Core Team, 2020).

## Results

We included 99,627 colon cancer cases (49.3% female) and 29,743 rectal cancer cases (41.3% female) aged 50-99 years old. Overall, 37.9% of colon cancer cases were diagnosed with localized disease, 35.6% with regional, and 22.7% with distant cancers. 4.2% of cases had unknown stage. The respective percentages for rectal cancer were 42.5%, 33.7%, 17.1% and 6.6%.

The distribution of stage by age at diagnosis for both colon and rectal cancers is shown in Figure 1. The percentage of missing stage was highest for the oldest age group in both cancer types, and it was higher in rectal cancer compared to colon cancer across all age groups.

**Figure 1.**
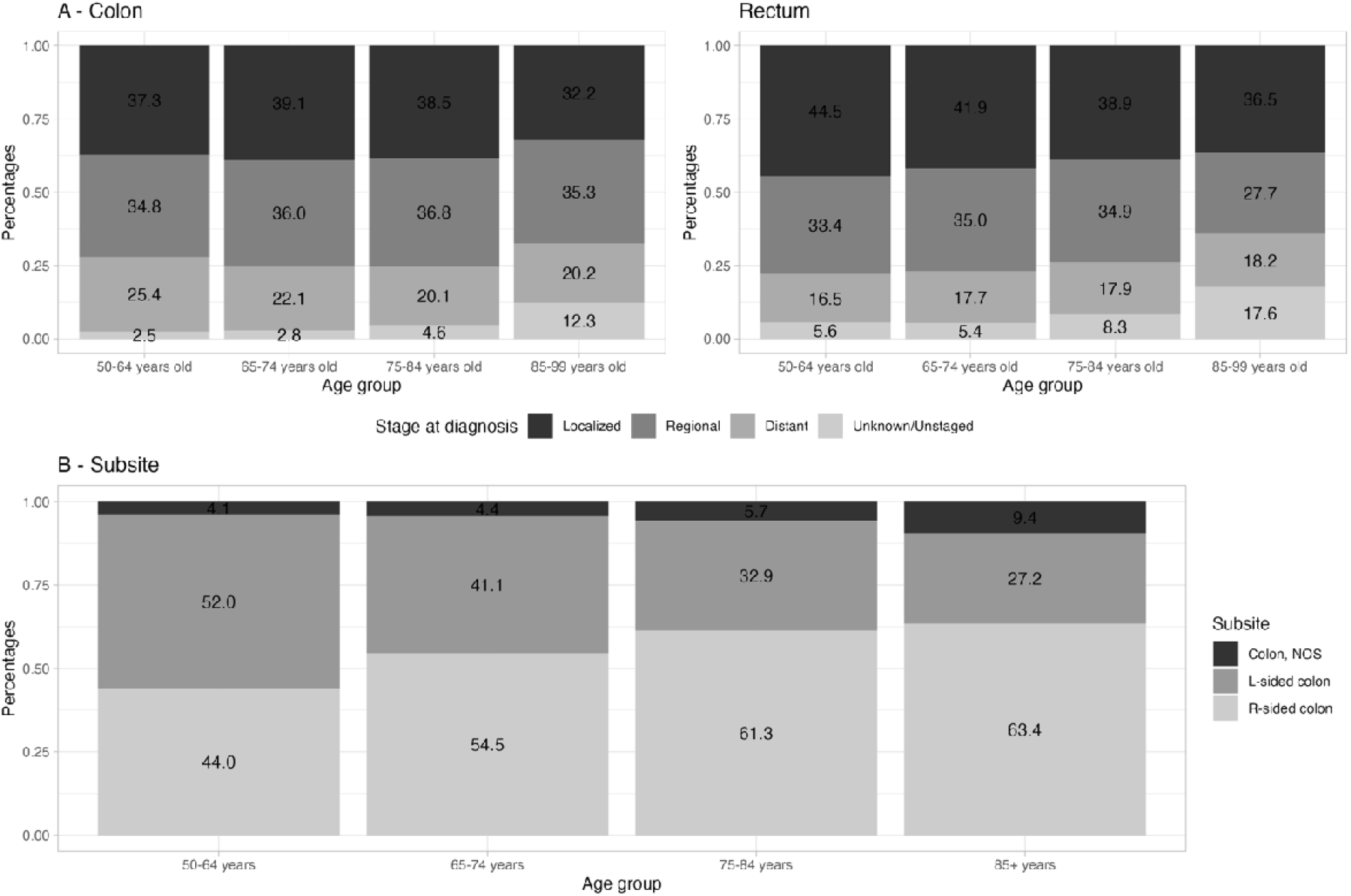
Distribution of (A) stage at diagnosis for colon and rectal cancer and (B) colon cancer sites, both by age group.

Most colon cancers were right-sided (53%). As age at diagnosis increased, the proportion of cancers right-sided tumours increased (Figure 1).

### One-year relative survival

Table 1 presents 1-year RS estimates by age and differences in 1-year RS between 50–64-year-old group and 75-84-year-old group for colon and rectal cancers, overall and by sex.

**Table 1.** One-year relative survival (RS) for colon and rectal cancers, respectively by age group, and sex

| Site | Age group (years) | All |  | Males |  | Females |  |
| --- | --- | --- | --- | --- | --- | --- | --- |
|  |  | n | RS (95%CI) | n | RS (95%CI) | n | RS (95%CI) |
| Colon | 50-64 | 37789 | 87.8 (87.5-88.2) | 21356 | 87.2 (86.7-87.6) | 16433 | 88.7 (88.2-89.2) |
|  | 65-74 | 28314 | 84.4 (83.9-84.8) | 15324 | 84.0 (83.4-84.6) | 12990 | 84.8 (84.1-85.4) |
|  | 75-84 | 21817 | 75.9 (75.3-76.5) | 9775 | 75.4 (74.4-76.3) | 12042 | 76.3 (75.5-77.1) |
|  | 85-99 | 11707 | 62.3 (61.3-63.3) | 4029 | 62.8 (60.9-64.5) | 7678 | 62.1 (60.8-63.3) |
|  | *Difference<sup>a</sup>* |  | *11.9 (11.2-12.7)* |  | *11.7 (10.7-12.8)* |  | *12.4 (11.5-13.4)* |
| Rectum | 50-64 | 16043 | 92.3 (91.8-92.7) | 9785 | 91.6 (91.0-92.2) | 6258 | 93.3 (92.6-93.9) |
|  | 65-74 | 7653 | 87.6 (86.8-88.4) | 4689 | 87.0 (85.9-88.0) | 2964 | 88.6 (87.3-89.7) |
|  | 75-84 | 4233 | 78.9 (77.5-80.3) | 2278 | 79.6 (77.7-81.4) | 1955 | 78.2 (76.1-80.1) |
|  | 85-99 | 1814 | 65.0 (62.3-67.5) | 715 | 68.5 (64.0-72.5) | 1099 | 62.7 (59.3-65.9) |
|  | *Difference<sup>a</sup>* |  | *13.3 (11.9-14.8)* |  | *12.0 (10.1-14.0)* |  | *15.1 (13.1-17.3)* |

<sup>a</sup> Absolute differences in RS between the 50-64 years age group and 75-84 years age group. CI: Confidence interval

For both colon and rectal cancers, the estimated 1-year RS decreased as age increased, with differences of around 12 to 13 %-points between 50-64 age group and 75-84 age group. One-year RS estimates for rectal cancer were slightly higher than those for colon cancer. For rectal cancer, females had slightly greater age-related differences than males (15.1 vs 12.0 %-points, respectively).

### By stage

Table 2 shows 1-year RS estimates by age, and differences in 1-year RS between the 50–64-year-old and 75-84-year-old groups stratified by stage at diagnosis, and sex. For both colon and rectal cancers, age-related differences in RS increased with increasing stage, and the magnitude of differences was similar for both types, ranging from 6 %-points in localized disease to 27 %-points in distant stage disease. Colon and rectal cancers with unknown stage had highest differences (37 and 28 %-points, respectively). Age-related differences in stage-specific RS for colon cancer were similar in males and females for all but distant stage tumours, where the age-related differences were higher in females than in males (31 vs. 24 %-points). For rectal cancer, females had greater age-related differences for distant and unknown stage cancers than males (29 and 36 %-points vs. 24 and 22 %-points, respectively).

**Table 2.**
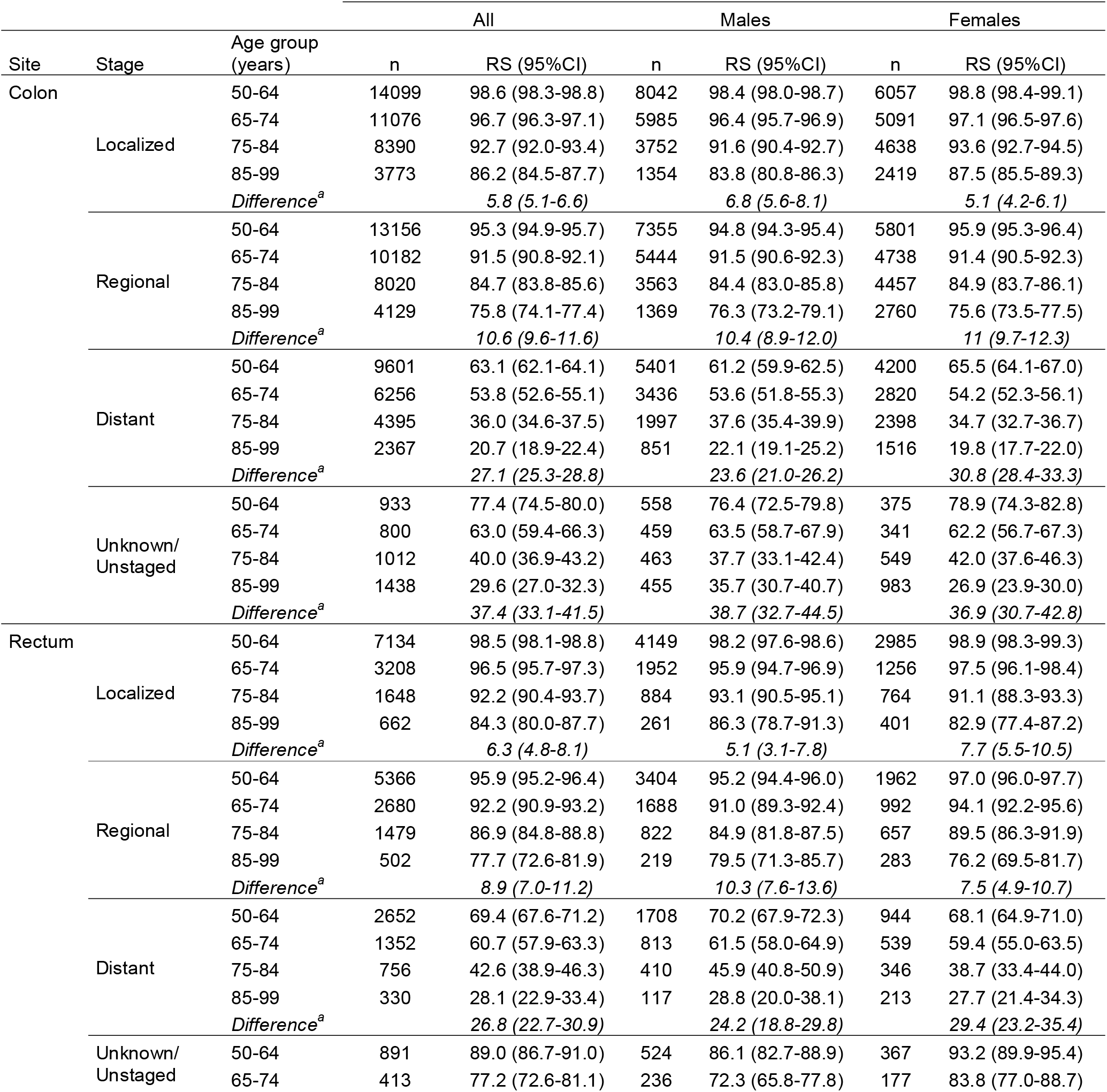
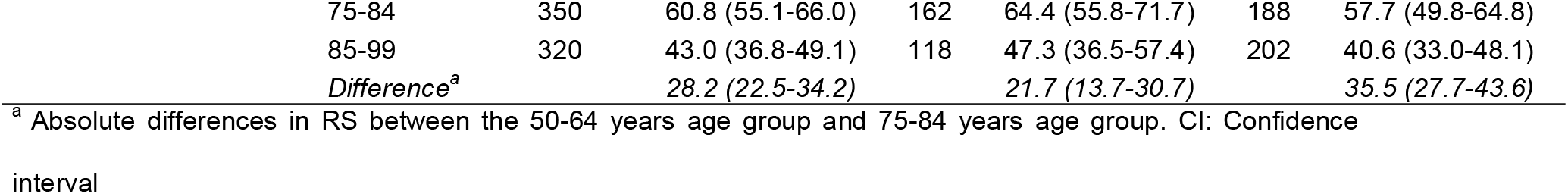
One-year relative survival (RS) for colon and rectum cancers, respectively, by age group, stage at diagnosis and sex

| Site | Stage | Age group (years) | All |  | Males |  | Females |  |
| --- | --- | --- | --- | --- | --- | --- | --- | --- |
|  |  |  | n | RS (95%CI) | n | RS (95%CI) | n | RS (95%CI) |
| Colon | Localized | 50-64 | 14099 | 98.6 (98.3-98.8) | 8042 | 98.4 (98.0-98.7) | 6057 | 98.8 (98.4-99.1) |
|  |  | 65-74 | 11076 | 96.7 (96.3-97.1) | 5985 | 96.4 (95.7-96.9) | 5091 | 97.1 (96.5-97.6) |
|  |  | 75-84 | 8390 | 92.7 (92.0-93.4) | 3752 | 91.6 (90.4-92.7) | 4638 | 93.6 (92.7-94.5) |
|  |  | 85-99 | 3773 | 86.2 (84.5-87.7) | 1354 | 83.8 (80.8-86.3) | 2419 | 87.5 (85.5-89.3) |
|  |  | *Difference<sup>a</sup>* |  | *5.8 (5.1-6.6)* |  | *6.8 (5.6-8.1)* |  | *5.1 (4.2-6.1)* |
|  | Regional | 50-64 | 13156 | 95.3 (94.9-95.7) | 7355 | 94.8 (94.3-95.4) | 5801 | 95.9 (95.3-96.4) |
|  |  | 65-74 | 10182 | 91.5 (90.8-92.1) | 5444 | 91.5 (90.6-92.3) | 4738 | 91.4 (90.5-92.3) |
|  |  | 75-84 | 8020 | 84.7 (83.8-85.6) | 3563 | 84.4 (83.0-85.8) | 4457 | 84.9 (83.7-86.1) |
|  |  | 85-99 | 4129 | 75.8 (74.1-77.4) | 1369 | 76.3 (73.2-79.1) | 2760 | 75.6 (73.5-77.5) |
|  |  | *Difference<sup>a</sup>* |  | *10.6 (9.6-11.6)* |  | *10.4 (8.9-12.0)* |  | *11 (9.7-12.3)* |
|  | Distant | 50-64 | 9601 | 63.1 (62.1-64.1) | 5401 | 61.2 (59.9-62.5) | 4200 | 65.5 (64.1-67.0) |
|  |  | 65-74 | 6256 | 53.8 (52.6-55.1) | 3436 | 53.6 (51.8-55.3) | 2820 | 54.2 (52.3-56.1) |
|  |  | 75-84 | 4395 | 36.0 (34.6-37.5) | 1997 | 37.6 (35.4-39.9) | 2398 | 34.7 (32.7-36.7) |
|  |  | 85-99 | 2367 | 20.7 (18.9-22.4) | 851 | 22.1 (19.1-25.2) | 1516 | 19.8 (17.7-22.0) |
|  |  | *Difference<sup>a</sup>* |  | *27.1 (25.3-28.8)* |  | *23.6 (21.0-26.2)* |  | *30.8 (28.4-33.3)* |
|  | Unknown/<br>Unstaged | 50-64 | 933 | 77.4 (74.5-80.0) | 558 | 76.4 (72.5-79.8) | 375 | 78.9 (74.3-82.8) |
|  |  | 65-74 | 800 | 63.0 (59.4-66.3) | 459 | 63.5 (58.7-67.9) | 341 | 62.2 (56.7-67.3) |
|  |  | 75-84 | 1012 | 40.0 (36.9-43.2) | 463 | 37.7 (33.1-42.4) | 549 | 42.0 (37.6-46.3) |
|  |  | 85-99 | 1438 | 29.6 (27.0-32.3) | 455 | 35.7 (30.7-40.7) | 983 | 26.9 (23.9-30.0) |
|  |  | *Difference<sup>a</sup>* |  | *37.4 (33.1-41.5)* |  | *38.7 (32.7-44.5)* |  | *36.9 (30.7-42.8)* |
| Rectum | Localized | 50-64 | 7134 | 98.5 (98.1-98.8) | 4149 | 98.2 (97.6-98.6) | 2985 | 98.9 (98.3-99.3) |
|  |  | 65-74 | 3208 | 96.5 (95.7-97.3) | 1952 | 95.9 (94.7-96.9) | 1256 | 97.5 (96.1-98.4) |
|  |  | 75-84 | 1648 | 92.2 (90.4-93.7) | 884 | 93.1 (90.5-95.1) | 764 | 91.1 (88.3-93.3) |
|  |  | 85-99 | 662 | 84.3 (80.0-87.7) | 261 | 86.3 (78.7-91.3) | 401 | 82.9 (77.4-87.2) |
|  |  | *Difference<sup>a</sup>* |  | *6.3 (4.8-8.1)* |  | *5.1 (3.1-7.8)* |  | *7.7 (5.5-10.5)* |
|  | Regional | 50-64 | 5366 | 95.9 (95.2-96.4) | 3404 | 95.2 (94.4-96.0) | 1962 | 97.0 (96.0-97.7) |
|  |  | 65-74 | 2680 | 92.2 (90.9-93.2) | 1688 | 91.0 (89.3-92.4) | 992 | 94.1 (92.2-95.6) |
|  |  | 75-84 | 1479 | 86.9 (84.8-88.8) | 822 | 84.9 (81.8-87.5) | 657 | 89.5 (86.3-91.9) |
|  |  | 85-99 | 502 | 77.7 (72.6-81.9) | 219 | 79.5 (71.3-85.7) | 283 | 76.2 (69.5-81.7) |
|  |  | *Difference<sup>a</sup>* |  | *8.9 (7.0-11.2)* |  | *10.3 (7.6-13.6)* |  | *7.5 (4.9-10.7)* |
|  | Distant | 50-64 | 2652 | 69.4 (67.6-71.2) | 1708 | 70.2 (67.9-72.3) | 944 | 68.1 (64.9-71.0) |
|  |  | 65-74 | 1352 | 60.7 (57.9-63.3) | 813 | 61.5 (58.0-64.9) | 539 | 59.4 (55.0-63.5) |
|  |  | 75-84 | 756 | 42.6 (38.9-46.3) | 410 | 45.9 (40.8-50.9) | 346 | 38.7 (33.4-44.0) |
|  |  | 85-99 | 330 | 28.1 (22.9-33.4) | 117 | 28.8 (20.0-38.1) | 213 | 27.7 (21.4-34.3) |
|  |  | *Difference<sup>a</sup>* |  | *26.8 (22.7-30.9)* |  | *24.2 (18.8-29.8)* |  | *29.4 (23.2-35.4)* |
|  | Unknown/<br>Unstaged | 50-64 | 891 | 89.0 (86.7-91.0) | 524 | 86.1 (82.7-88.9) | 367 | 93.2 (89.9-95.4) |
|  |  | 65-74 | 413 | 77.2 (72.6-81.1) | 236 | 72.3 (65.8-77.8) | 177 | 83.8 (77.0-88.7) |

|  |  |  |  |  |  |  |
| --- | --- | --- | --- | --- | --- | --- |
| 75-84 | 350 | 60.8 (55.1-66.0) | 162 | 64.4 (55.8-71.7) | 188 | 57.7 (49.8-64.8) |
| 85-99 | 320 | 43.0 (36.8-49.1) | 118 | 47.3 (36.5-57.4) | 202 | 40.6 (33.0-48.1) |
| *Difference<sup>a</sup>* |  | *28.2 (22.5-34.2)* |  | *21.7 (13.7-30.7)* |  | *35.5 (27.7-43.6)* |

<sup>a</sup> Absolute differences in RS between the 50-64 years age group and 75-84 years age group. CI: Confidence interval

### Colon, by tumour site

Supplementary Table 1 presents age-specific one-year RS for colon sites by sex. Age-related differences in RS were greatest for tumours of the colon, not otherwise specified (NOS)’ (25 %-points) and smallest for tumours on the right-side of the colon (8 %-points, supplementary Table 1). Differences were similar between sexes.

Figure 2 and supplementary Table 2 show age-related differences in RS stratified by tumour site, stage at diagnosis, and sex. There was a pattern of increasing age-related differences with increasing stage for all sites except ‘colon, NOS’ overall and when stratified by sex. Differences in RS were highest for cancers of unknown stage. Age-related differences in one-year RS were higher in females than in males with distant cancer across all sites (28 vs 18 %-points for right-sided colon cancer, 32 vs 26 %-points for left-sided colon cancer, and 24 vs 19 %-points for colon, NOS).

**Figure 2.**
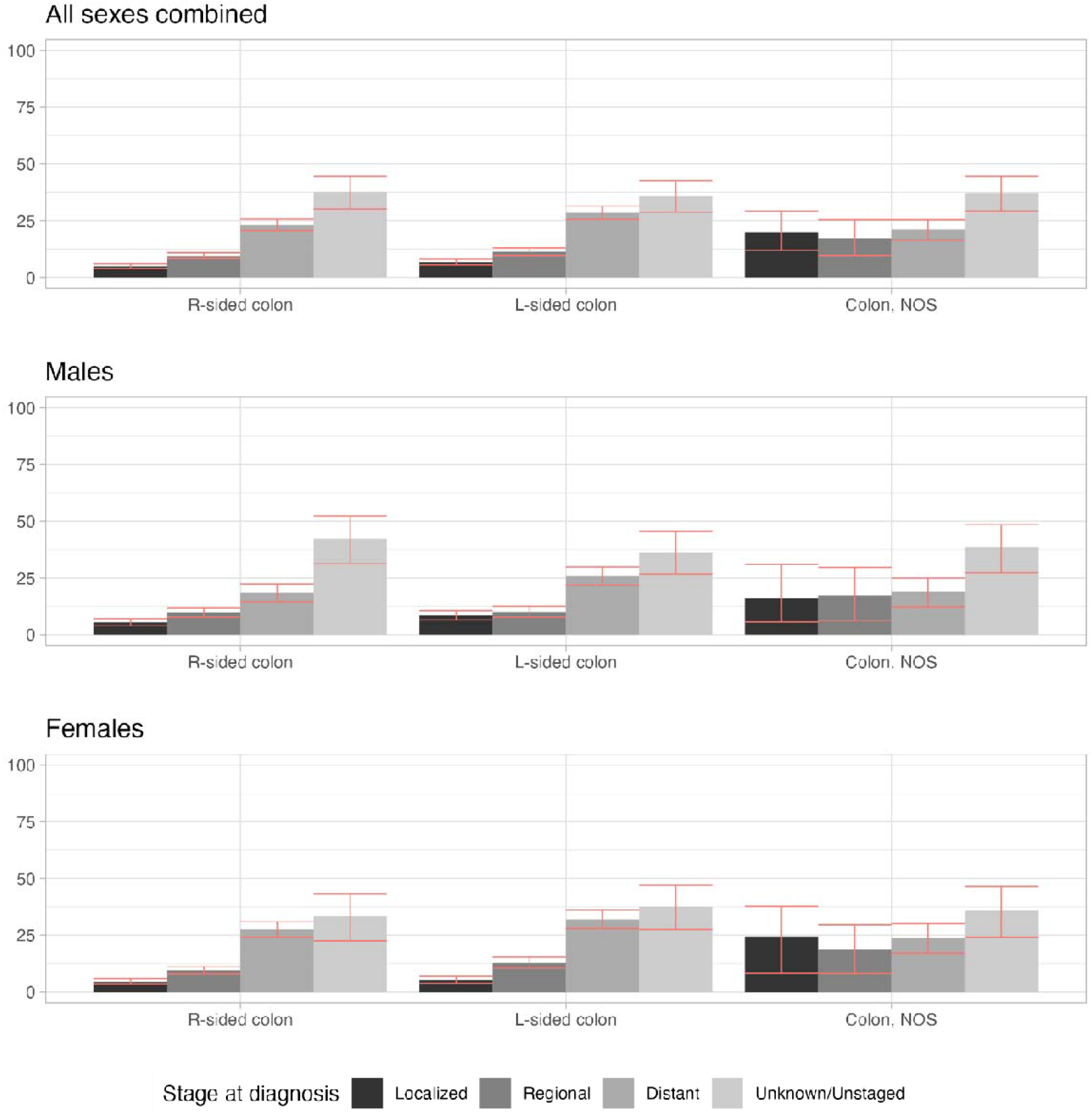
Absolute difference in one-year relative survival from colon cancer between 50–64-year-olds and 75-84-year-olds by tumour sites, stage at diagnosis, and sex. Note. Red bars represent 95% confidence intervals.

### Colon, by histology

Overall, the smallest age-related differences in RS were observed for signet ring cell carcinomas (7 %-points) and the greatest difference was seen for ‘other’ histology (42 %-points, Supplementary Table 3). Differences were 10%-points for adenocarcinomas and mucinous adenocarcinomas. Age-related differences increased with increasing stage for all histologies (supplementary Table 3).

### One-year relative surviving conditioning on surviving one year of diagnosis

In patients who survived the first year after diagnosis, age-related differences in 1-year relative survival were greatly reduced in both cancer types and all stages (Appendix, Table 2) but remained important for distant and unstaged cancers.

## Discussion

Using the most-up-to-date relative survival estimates from 18 population-based cancer registries in the US, we confirmed poorer one-year relative survival for colon and rectal cancers in older adults compared to middle-aged adults. With few exceptions, age-related differences were consistent by tumour site, histology, and sex and consistently worsened with increasing stage of disease. Because age-related differences in survival were observed for all histologies and topographies, overall age-related differences were not driven by differences in the distribution of tumour site or histology by age, or by large age-related differences in survival in specific tumour types or histologies.

The persistent poorer relative survival from colon and rectal cancers among older adults we have reported is congruent with findings from the International Cancer Benchmarking Partnership (ICBP), which estimated one-year relative survival for both types in people under and over 75 in seven high-income countries with different healthcare systems and health coverage than in the US.^3^

Consistent with our findings, increasing age-related differences in colon and rectal cancer survival with worsening stage of diagnosis and large differences for unstaged diseases have been observed in ICBP and other countries,^5,3,13^ which have also been shown for other cancer types in the US.^19^

Regarding sex, the greater age-related differences in colon cancer survival in females with distant disease than males were also observed in New Zealand and Finland.^5,20^ Some authors have suggested a possible involvement of sex hormones, but no causal mechanisms have been identified so far.^21^ As such, variation in age differences in colon and rectal survival by sex merit further investigation.

With respect to colon, few studies have provided relative survival estimates stratified by both age group and tumour site. Two Korean studies also found relatively consistent age-related differences in colon cancer survival by tumour site, but differed by finding larger age-related differences for rectal than colon cancer.^22,23^ In Finland, age-related differences in 1-year relative survival appeared consistent for tumour sites of colon cancers, but were notably higher for tumours whose topography was classified as “other”.^20^

Finally, with respect to histology of colon cancer, while the Finish study separated ‘carcinoid tumours of the colon’ from ‘carcinoma of the colon’, little could be interpreted given small numbers of cases within each age group.^20^

Stage and treatment are determinants of cancer survival. Age-related differences in survival within stage categories suggest that there are differences in treatment strategies due to either medical consideration and/or patients’ preferences, and/or differences in post-treatment mortality.

In the case of colorectal cancer, surgery is the mainstay treatment for non-metastatic colorectal tumours, including in older adults.^24,25^ While age-specific surgery rates are not documented at population-level in the US, a European study showed that in high-income countries, adults aged over 75 had consistently lower surgical resection rates than younger adults.^26^ This age-related difference in surgery rates increased with increasing stage and was greater for rectal cancer.

Several reasons may explain lower surgery rates in older adults. First, it has been shown that older adults, even when eligible for surgery, are more likely to refuse it and those that refuse had poorer survival.^27^ Second, frail older patients may not be considered eligible for surgery because of higher risk of complications (see below).^28,29^

Previous studies have suggested that age-related differences in colon and rectal cancer survival arise in the first year after diagnosis.^4,30,31^ This is consistent with the reduction in age-related differences we observed when conditioning on surviving on the first year after diagnosis. Older patients have higher post-operative mortality rates after elective surgery; however, recent evidence in European countries showed that the gap in post-operative complication rates between middle-aged and older patients is narrowing thanks to recent changes in management, better pre-operative assessment, and rehabilitation.^28^ In Denmark, increased use of major surgical resection in patients aged 80 and over was associated with higher 30-day post-operative mortality, but also associated with improved one-year survival, suggesting that a trade-off between short and long term risks may need to be considered.^32^ Furthermore, older adults are more likely to present as an emergency (e.g., with a bowel obstruction), which is associated with poorer outcomes.^33^

We observed consistently higher age-related differences for cancers of unknown stage or topography. From available SEER data, we cannot distinguish to what extent missingness is due to the cancer stage or topography being truly unknown or if these features were not reported to the registry but known by the clinician. Cancers may be clinically unstaged or of unknown topography due to patients’ preferences (e.g., patients refusing full diagnostic work-up) or patients’ health status being too frail to undergo full work-up. If more missingness among older adults was attributable to these reasons, this may explain the higher age-related differences within the unstaged category. The extent to which errors or omissions in reporting of known stage are differential by age is not known. Age has been shown to be a determinant of missingness in administrative data, but an understanding of how the causes of missingness differ by age is necessary to fully interpret these findings.^34^ As age-related differences in survival were higher in advanced staged disease, they could theoretically be minimized by a) decreasing the proportion of cancers diagnosed among older adults at advanced stage and/or b) more effectively treating older people with more advanced disease. Ideally, efforts to reduce advanced stage cancer diagnoses will benefit people of all ages, and if stage distribution can be shifted toward more localized/regional disease and fewer emergency presentations, we will observe narrower gaps in survival between older and younger adults. A recent systematic review showed that multiple factors influence the patient and general practitioners’ decision to investigate cancer symptoms in older adults, including the presence of frailty, comorbidities, cognitive impairment, family, and carer involvement; the most important factor was the lack of consultation time. However, the review did not compare age groups, potentially reflecting limited evidence of how and whether these factors differ by age.^35^ Furthermore, improving treatment outcomes by properly selecting older adults who will benefit from curative treatment, for instance by using a comprehensive geriatric assessment, may increase survival probabilities for older adults. While patient preference to refuse treatment may play a part in reduced survival in older adults, our data was insufficient to determine to what extent this was the case. Previous research that has examined patient preference and refusal of cancer treatment in older adults has reported that older adults (as younger adults) prioritize quality of life and overall survival.^36^ The most consistent determinant of refusal or acceptation of treatment was physician recommendation.^37^ Further research on this would be useful to better interpret age-related disparities. Although fully eliminating the survival gap between middle-aged and older adults may be utopian, improving outcomes for older adults is worthwhile and realistic.

Our study has limitations. First, we regret the lack of data on other factors that may influence cancer management, and ultimately, survival, such as geriatric conditions (e.g., frailty, falls), comorbidity, social factors (e.g., social support), among others. Second, the estimation of relative survival relies on background mortality rates obtained from lifetables and implies that in the absence of cancer, cancer patients would have the same mortality risk as the general population. While the US life tables account for differences in age, sex, ethnicity and county SES between cancer patients and the general population, they are not specific to other factors influencing prognosis and mortality (such as comorbidities), for which the prevalence may differ between cancer patients and the general population. If life tables are differentially biased (e.g. by age and comorbidity), our estimates of age-related differences in RS may therefore be underestimated.^38^

The strengths of the study include the description of age-related disparities by tumour site and histology, the coverage of 30% of the US population, and the high quality of SEER data.

## Conclusion

We showed persistent differences in colon and rectal cancer survival in relation to age, especially during the initial year, which are unlikely to be driven by differences in histology or tumour site. Further research is warranted to disentangle the share of these differences that are avoidable through improved diagnosis and/or treatment. Integrating more detailed information on geriatric conditions to cancer registries, or detailed linkage studies may help to achieve this.

## Data Availability

SEER data and cancer survival estimates are publicly available and were retrieved through SEER*Stat software version 8.3.9 (National Cancer Institute - https://seer.cancer.gov/seerstat/).

## Acknowledgement

Authors thank Dr Hadrien Charvat for his help with calculating 95% confidence interval around differences in relative survival.

