## Supplemental material for "Age-related differences in colon and rectal cancer survival: An analysis of United States SEER-18 data"

**Supplementary Materials**

**Supplementary Table 1**. One-year relative survival (RS) for colon cancer subsites by age group and sex

|  |  | **All sexes combined** | | **Males** | | **Females** | |
| --- | --- | --- | --- | --- | --- | --- | --- |
|  | **Age group**  **(years)** | **n** | **RS (95%CI)** | **n** | **RS (95%CI)** | **n** | **RS (95%CI)** |
| Right-sided colon | 50-64 | 16618 | 87.2 (86.6-87.7) | 8852 | 86.5 (85.7-87.2) | 7766 | 88.0 (87.2-88.7) |
|  | 65-74 | 15443 | 86.1 (85.5-86.6) | 7659 | 85.9 (85.0-86.7) | 7784 | 86.3 (85.4-87.1) |
|  | 75-84 | 13383 | 79.4 (78.6-80.2) | 5513 | 78.8 (77.5-80.0) | 7870 | 79.9 (78.9-80.8) |
|  | 85-99 | 7422 | 68.4 (67.1-69.6) | 2381 | 68.3 (65.9-70.6) | 5041 | 68.4 (66.9-69.9) |
|  | *Difference* |  | *7.7 (6.8-8.7)* |  | *7.7 (6.3-9.1)* |  | *8.1 (6.9-9.3)* |
| Left-sided colon | 50-64 | 19636 | 90.6 (90.2-91.1) | 11608 | 89.8 (89.3-90.4) | 8028 | 91.8 (91.1-92.4) |
|  | 65-74 | 11628 | 85.9 (85.2-86.5) | 6959 | 85.6 (84.7-86.5) | 4669 | 86.2 (85.1-87.2) |
|  | 75-84 | 7182 | 76.6 (75.5-77.7) | 3709 | 76.4 (74.8-77.9) | 3473 | 76.8 (75.2-78.3) |
|  | 85-99 | 3182 | 61.7 (59.7-63.6) | 1292 | 62.6 (59.4-65.6) | 1890 | 61.1 (58.5-63.6) |
|  | *Difference* |  | *14.0 (12.9-15.2)* |  | *13.4 (11.9-15.1)* |  | *15.0 (13.4-16.7)* |
| Colon, not otherwise specified | 50-64 | 1535 | 59.2 (56.7-61.7) | 896 | 58.6 (55.3-61.8) | 639 | 60.0 (56.0-63.8) |
|  | 65-74 | 1243 | 49.6 (46.7-52.4) | 706 | 48.4 (44.5-52.1) | 537 | 51.2 (46.8-55.4) |
|  | 75-84 | 1252 | 34.1 (31.3-36.8) | 553 | 34.7 (30.6-38.9) | 699 | 33.5 (29.9-37.2) |
|  | 85-99 | 1103 | 23.2 (20.6-26.0) | 356 | 26.2 (21.3-31.4) | 747 | 21.9 (18.7-25.1) |
|  | *Difference* |  | *25.2 (21.4-28.8)* |  | *23.9 (18.6-29.2)* |  | *26.5 (21.1-31.7)* |

**Supplementary Table 2.** One-year relative survival (RS) by age groups and difference in RS between 50-64- and 75–84-year-old groups by topography, stage, and sex.

|  |  |  | **All sexes combined** | | **Males** | | **Females** | |
| --- | --- | --- | --- | --- | --- | --- | --- | --- |
| **Subsite** | **Stage** | **Age group**  **(years)** | **n** | **RS (95%CI)** | **n** | **RS (95%CI)** | **n** | **RS (95%CI)** |
| R-sided colon | Localized | 50-64 | 6082 | 98.3 (97.8-98.6) | 3294 | 98.1 (97.4-98.6) | 2788 | 98.4 (97.8-98.9) |
|  |  | 65-74 | 6266 | 96.8 (96.2-97.4) | 3112 | 96.5 (95.5-97.2) | 3154 | 97.2 (96.4-97.9) |
|  |  | 75-84 | 5489 | 93.4 (92.5-94.2) | 2272 | 92.7 (91.1-94.0) | 3217 | 93.9 (92.7-94.9) |
|  |  | 85-99 | 2662 | 88.3 (86.4-89.9) | 882 | 84.7 (80.9-87.8) | 1780 | 89.9 (87.6-91.8) |
|  |  | *Difference* |  | *4.9 (3.9-5.9)* |  | *5.4 (4-7.1)* |  | *4.5 (3.4-5.8)* |
|  | Regional | 50-64 | 6016 | 94.6 (94.0-95.2) | 3208 | 94.1 (93.1-94.9) | 2808 | 95.2 (94.3-96.0) |
|  |  | 65-74 | 5875 | 91.3 (90.4-92.0) | 2908 | 91.3 (90.0-92.4) | 2967 | 91.3 (90.1-92.3) |
|  |  | 75-84 | 5270 | 85.1 (84.0-86.2) | 2125 | 84.3 (82.3-86.0) | 3145 | 85.7 (84.2-87.0) |
|  |  | 85-99 | 2908 | 76.3 (74.3-78.2) | 886 | 78.7 (74.8-82.0) | 2022 | 75.3 (72.9-77.5) |
|  |  | *Difference* |  | *9.5 (8.3-10.8)* |  | *9.8 (7.8-11.9)* |  | *9.6 (7.9-11.2)* |
|  | Distant | 50-64 | 4263 | 61.6 (60.1-63.1) | 2211 | 58.8 (56.6-60.8) | 2052 | 64.7 (62.6-66.8) |
|  |  | 65-74 | 3046 | 55.5 (53.6-57.3) | 1502 | 55.1 (52.5-57.6) | 1544 | 55.9 (53.3-58.3) |
|  |  | 75-84 | 2238 | 38.5 (36.4-40.6) | 962 | 40.4 (37.1-43.6) | 1276 | 37.2 (34.4-39.9) |
|  |  | 85-99 | 1253 | 23.2 (20.7-25.8) | 423 | 23.8 (19.4-28.4) | 830 | 22.9 (19.9-26.1) |
|  |  | *Difference* |  | *23.1 (20.6-25.7)* |  | *18.4 (14.6-22.3)* |  | *27.6 (24.1-31.0)* |
|  | Unknown/unstaged | 50-64 | 257 | 79.6 (74.0-84.2) | 139 | 81.8 (74.0-87.4) | 118 | 77.1 (68.2-83.8) |
|  |  | 65-74 | 256 | 67.7 (61.4-73.2) | 137 | 67.7 (58.7-75.1) | 119 | 67.7 (58.2-75.5) |
|  |  | 75-84 | 386 | 41.9 (36.8-47.0) | 154 | 39.4 (31.3-47.5) | 232 | 43.6 (36.9-50.1) |
|  |  | 85-99 | 599 | 35.8 (31.5-40.2) | 190 | 42.7 (34.5-50.8) | 409 | 32.6 (27.6-37.7) |
|  |  | *Difference* |  | *37.7 (30.1-44.6)* |  | *42.3 (31.3-52.3)* |  | *33.5 (22.5-43.1)* |
| L-sided colon | Localized | 50-64 | 7827 | 98.8 (98.4-99.1) | 4629 | 98.6 (98.1-99.0) | 3198 | 99.0 (98.5-99.3) |
|  |  | 65-74 | 4643 | 96.6 (95.9-97.2) | 2781 | 96.3 (95.3-97.1) | 1862 | 97.0 (95.9-97.8) |
|  |  | 75-84 | 2785 | 92.1 (90.7-93.3) | 1428 | 90.3 (88.1-92.0) | 1357 | 93.8 (91.9-95.2) |
|  |  | 85-99 | 1068 | 80.9 (77.6-83.8) | 456 | 81.8 (76.4-86.1) | 612 | 80.2 (75.8-83.9) |
|  |  | *Difference* |  | *6.7 (5.5-8.1)* |  | *8.4 (6.5-10.5)* |  | *5.2 (3.7-7.1)* |
|  | Regional | 50-64 | 6875 | 96.2 (95.6-96.6) | 3989 | 95.8 (95.0-96.4) | 2886 | 96.7 (95.9-97.3) |
|  |  | 65-74 | 4104 | 92.1 (91.2-93.0) | 2427 | 92.0 (90.7-93.2) | 1677 | 92.3 (90.8-93.6) |
|  |  | 75-84 | 2552 | 84.9 (83.2-86.4) | 1345 | 85.7 (83.4-87.8) | 1207 | 83.9 (81.4-86.1) |
|  |  | 85-99 | 1089 | 75.7 (72.3-78.7) | 432 | 74.1 (68.5-78.9) | 657 | 76.7 (72.4-80.4) |
|  |  | *Difference* |  | *11.3 (9.7-13.0)* |  | *10.0 (7.8-12.5)* |  | *12.8 (10.5-15.4)* |
|  | Distant | 50-64 | 4546 | 69.0 (67.6-70.3) | 2742 | 67.4 (65.6-69.2) | 1804 | 71.3 (69.1-73.3) |
|  |  | 65-74 | 2553 | 58.5 (56.5-60.4) | 1556 | 58.4 (55.9-60.9) | 997 | 58.6 (55.4-61.7) |
|  |  | 75-84 | 1528 | 40.4 (37.9-43.0) | 767 | 41.6 (37.9-45.2) | 761 | 39.3 (35.7-42.8) |
|  |  | 85-99 | 662 | 23.1 (19.7-26.6) | 280 | 24.6 (19.3-30.2) | 382 | 22.0 (17.7-26.6) |
|  |  | *Difference* |  | *28.5 (25.6-31.4)* |  | *25.8 (21.8-29.9)* |  | *32.0 (27.9-36.2)* |
|  | Unknown/unstaged | 50-64 | 388 | 84.3 (80.1-87.6) | 248 | 80.1 (74.3-84.7) | 140 | 91.5 (85.3-95.2) |
|  |  | 65-74 | 328 | 69.7 (64.2-74.5) | 195 | 72.4 (65.2-78.4) | 133 | 65.7 (56.6-73.3) |
|  |  | 75-84 | 317 | 48.5 (42.6-54.2) | 169 | 43.8 (35.8-51.5) | 148 | 54.0 (45.1-62.1) |
|  |  | 85-99 | 363 | 34.1 (28.7-39.5) | 124 | 38.2 (28.7-47.7) | 239 | 31.9 (25.5-38.5) |
|  |  | *Difference* |  | *35.7 (28.8-42.6)* |  | *36.2 (26.8-45.5)* |  | *37.5 (27.5-47.1)* |
| Colon,  Not otherwise specified | Localized | 50-64 | 190 | 98.4 (94.2-99.6) | 119 | 98.0 (91.8-99.5) | 71 | 99.0 (85.0-99.9) |
|  |  | 65-74 | 167 | 94.6 (88.8-97.4) | 92 | 93.3 (84.2-97.2) | 75 | 95.3 (86.3-98.5) |
|  |  | 75-84 | 116 | 78.6 (69.0-85.5) | 52 | 82.0 (66.8-90.7) | 64 | 74.6 (60.7-84.2) |
|  |  | 85-99 | 43 | 82.6 (63.5-92.3) | 16 | 81.5 (43.6-95.1) | 27 | 83.0 (53.9-94.6) |
|  |  | *Difference* |  | *19.9 (11.9-29.3)* |  | *16.0 (5.6-31.0)* |  | *24.4 (8.2-37.7)* |
|  | Regional | 50-64 | 265 | 89.5 (84.8-92.8) | 158 | 87.1 (80.4-91.7) | 107 | 92.9 (85.5-96.6) |
|  |  | 65-74 | 203 | 84.2 (77.9-88.8) | 109 | 86.2 (77.3-91.8) | 94 | 81.8 (71.9-88.5) |
|  |  | 75-84 | 198 | 72.2 (64.7-78.3) | 93 | 69.8 (58.2-78.8) | 105 | 74.2 (63.8-82.1) |
|  |  | 85-99 | 132 | 64.8 (54.3-73.5) | 51 | 53.3 (36.1-67.7) | 81 | 72.0 (58.4-81.9) |
|  |  | *Difference* |  | *17.3 (9.7-25.4)* |  | *17.3 (6.2-29.7)* |  | *18.7 (8.1-29.7)* |
|  | Distant | 50-64 | 792 | 37.3 (33.9-40.7) | 448 | 35.2 (30.7-39.7) | 344 | 40.1 (34.8-45.2) |
|  |  | 65-74 | 657 | 28.2 (24.7-31.7) | 378 | 27.3 (22.9-32.0) | 279 | 29.3 (24.0-34.8) |
|  |  | 75-84 | 629 | 16.3 (13.5-19.4) | 268 | 16.4 (12.1-21.3) | 361 | 16.3 (12.6-20.4) |
|  |  | 85-99 | 452 | 10.1 (7.4-13.3) | 148 | 12.7 (7.6-19.2) | 304 | 8.9 (5.8-12.6) |
|  |  | *Difference* |  | *21.0 (16.4-25.4)* |  | *18.8 (12.3-25.0)* |  | *23.8 (17.1-30.2)* |
|  | Unknown/unstaged | 50-64 | 288 | 66.2 (60.3-71.5) | 171 | 66.8 (59.0-73.4) | 117 | 65.5 (55.9-73.4) |
|  |  | 65-74 | 216 | 47.3 (40.4-54.0) | 127 | 45.7 (36.6-54.2) | 89 | 49.7 (38.7-59.8) |
|  |  | 75-84 | 309 | 29.0 (23.8-34.3) | 140 | 28.4 (20.9-36.4) | 169 | 29.5 (22.5-36.7) |
|  |  | 85-99 | 476 | 18.6 (14.9-22.5) | 141 | 24.0 (16.6-32.0) | 335 | 16.3 (12.3-20.8) |
|  |  | *Difference* |  | *37.2 (29.3-44.6)* |  | *38.4 (27.3-48.5)* |  | *36.0 (24.1-46.6)* |

**Supplementary Table 3.** One-year relative survival (RS) by age groups and difference in RS between 50-64- and 75–84-year-old groups by histology and stage

|  |  | **Adenocarcinoma** | | **Mucinous adenocarcinomas** | | **Signet ring cell carcinoma** | | **Other** | |
| --- | --- | --- | --- | --- | --- | --- | --- | --- | --- |
| **Stage** | **Age group (years)** | **n** | **RS (95%CI)** | **n** | **RS (95%CI)** | **n** | **RS (95%CI)** | **n** | **RS (95%CI)** |
| All stages combined | 50-64 | 32692 | 88.5 (88.1-88.8) | 2648 | 89.3 (88.0-90.5) | 403 | 69.0 (64.1-73.3) | 2046 | 79.6 (77.7-81.3) |
|  | 65-74 | 24713 | 85.5 (85.0-85.9) | 2068 | 85.4 (83.7-87.0) | 329 | 65.0 (59.4-70.0) | 1204 | 65.4 (62.6-68.1) |
|  | 75-84 | 18787 | 77.9 (77.2-78.5) | 1759 | 79.2 (77.0-81.3) | 246 | 61.5 (54.7-67.6) | 1025 | 37.6 (34.5-40.7) |
|  | 85-99 | 9435 | 67.1 (65.9-68.2) | 899 | 74.9 (71.2-78.2) | 106 | 51.0 (39.8-61.2) | 1267 | 18.5 (16.2-20.9) |
|  | *Difference* |  | *10.6 (9.9-11.4)* |  | *10.1 (7.7-12.6)* |  | *7.4 (-0.3-15.5)* |  | *42.0 (38.4-45.5)* |
| Localized | 50-64 | 12594 | 98.5 (98.3-98.8) | 590 | 98.4 (96.7-99.3) | 38 | 97.5 (81.7-99.7) | 879 | 98.6 (97.2-99.3) |
|  | 65-74 | 10126 | 96.9 (96.4-97.3) | 553 | 94.3 (91.6-96.2) | 31 | 97.9 (8.9-100) | 359 | 95.8 (92.4-97.7) |
|  | 75-84 | 7649 | 92.8 (92.0-93.5) | 552 | 94.9 (91.6-96.9) | 16 | 90.4 (53.1-98.4) | 158 | 82.0 (74.1-87.7) |
|  | 85-99 | 3375 | 86.5 (84.8-88.1) | 279 | 89.9 (84.0-93.7) |  | -* | 103 | 59.4 (47.2-69.7) |
|  | *Difference* |  | *5.8 (5.0-6.6)* |  | *3.6 (1.0-6.9)* |  | *7.1 (-10.1-43.3)* |  | *16.6 (10.7-24.5)* |
| Regional | 50-64 | 11485 | 95.6 (95.2-96.0) | 1079 | 95.6 (94.0-96.7) | 154 | 83.5 (76.4-88.7) | 438 | 91.6 (88.4-94.0) |
|  | 65-74 | 8828 | 91.8 (91.1-92.4) | 939 | 91.9 (89.8-93.7) | 141 | 78.6 (70.5-84.8) | 274 | 86.9 (81.9-90.6) |
|  | 75-84 | 6873 | 85.4 (84.4-86.3) | 822 | 84.3 (81.2-87.0) | 123 | 71.6 (61.9-79.3) | 202 | 71.9 (64.6-78.0) |
|  | 85-99 | 3509 | 76.4 (74.6-78.1) | 441 | 81.4 (76.1-85.6) | 48 | 61.9 (44.2-75.4) | 131 | 44.9 (35.1-54.1) |
|  | *Difference* |  | *10.2 (9.2-11.3)* |  | *11.2 (8.2-14.6)* |  | *11.9 (1.5-22.9)* |  | *19.7 (12.9-27.4)* |
| Distant | 50-64 | 7942 | 63.1 (62.0-64.1) | 951 | 76.9 (74.0-79.5) | 207 | 52.3 (45.2-58.9) | 501 | 42.0 (37.6-46.3) |
|  | 65-74 | 5177 | 54.6 (53.2-56.0) | 555 | 65.8 (61.6-69.7) | 148 | 42.7 (34.5-50.6) | 376 | 29.5 (24.8-34.2) |
|  | 75-84 | 3583 | 37.4 (35.8-39.1) | 362 | 45.8 (40.4-51.1) | 89 | 37.4 (27.2-47.7) | 361 | 11.7 (8.6-15.4) |
|  | 85-99 | 1692 | 23.9 (21.8-26.2) | 170 | 33.8 (26.1-41.7) | 33 | 20.2 (8.1-36.1) | 472 | 4.0 (2.4-6.3) |
|  | *Difference* |  | *25.6 (23.7-27.6)* |  | *31.1 (25.1-37.1)* |  | *14.8 (2.5-27.0)* |  | *30.3 (24.6-35.6)* |
| Unknown/unstaged | 50-64 | 671 | 80.9 (77.6-83.8) | 28 | 79.3 (58.6-90.4) | 6 | 83.7 (26.6-97.6) | 228 | 66.7 (60.0-72.5) |
|  | 65-74 | 582 | 66.9 (62.8-70.7) | 21 | 82.4 (56.8-93.6) | 9 | 25.5 (3.5-57.2) | 195 | 48.8 (41.4-55.7) |
|  | 75-84 | 682 | 47.9 (43.8-51.8) | 23 | 49.3 (27.4-67.9) |  | -* | 304 | 22.3 (17.6-27.3) |
|  | 85-99 | 859 | 37.9 (34.2-41.5) | 9 | 24.3 (3.5-55.0) |  | -* | 561 | 17.2 (13.9-20.7) |
|  | *Difference* |  | *33.1 (28.0-38.0)* |  | *30.0 (2.6-54.7)* |  | *-** |  | *44.4 (36.1-51.8)* |

*CI: Confidence intervals; RS: relative survival*

* 0 cases

**Supplementary Table 4.** One-year relative survival (RS) conditioning on surviving one year and 95% confidence interval in colon and rectal cancers for all stages combined and by stage

|  |  | **Colon** | | **Rectum** | |
| --- | --- | --- | --- | --- | --- |
| **Stage** | **Age group (years)** | **n** | **RS (95%CI)** | **n** | **RS (95%CI)** |
| All | 50-64 | 31871 | 91.4 (91.1-91.8) | 14249 | 93.2 (92.7-93.6) |
|  | 65-74 | 22972 | 91.6 (91.1-92.0) | 6456 | 91.5 (90.7-92.3) |
|  | 75-84 | 15440 | 90.7 (90.0-91.2) | 3128 | 89.2 (87.7-90.5) |
|  | 85-99 | 6308 | 90.0 (88.7-91.2) | 1017 | 78.7 (75.0-81.9) |
|  | *Difference* |  | *0.8 (0.1-1.5)* |  | *4.0 (2.6-5.6)* |
| Localized | 50-64 | 13259 | 99.1 (98.9-99.3) | 6723 | 98.8 (98.4-99.1) |
|  | 65-74 | 10299 | 98.7 (98.2-99.0) | 2977 | 98.1 (97.2-98.7) |
|  | 75-84 | 7251 | 98.1 (97.3-98.6) | 1427 | 96.5 (94.5-97.8) |
|  | 85-99 | 2795 | 99.3 (96.5-99.9) | 484 | 89.4 (84.0-93.1) |
|  | *Difference* |  | *1.1 (0.5-1.9)* |  | *2.3 (1.0-4.4)* |
| Regional | 50-64 | 12022 | 94.7 (94.2-95.1) | 4984 | 94.7 (93.9-95.4) |
|  | 65-74 | 8939 | 93.8 (93.2-94.4) | 2380 | 92.1 (90.7-93.3) |
|  | 75-84 | 6322 | 90.6 (89.6-91.6) | 1206 | 91.2 (88.7-93.1) |
|  | 85-99 | 2724 | 90.6 (88.5-92.3) | 338 | 80.5 (73.7-85.7) |
|  | *Difference* |  | *4.0 (3.0-5.1)* |  | *3.5 (1.5-6.0)* |
| Distant | 50-64 | 5908 | 67.8 (66.5-69.1) | 1792 | 67.1 (64.6-69.3) |
|  | 65-74 | 3255 | 64.9 (63.1-66.6) | 797 | 67.2 (63.4-70.6) |
|  | 75-84 | 1490 | 57.8 (55.0-60.6) | 297 | 53.9 (47.4-59.9) |
|  | 85-99 | 428 | 49.5 (43.6-55.1) | 80 | 49.0 (35.7-61.1) |
|  | *Difference* |  | *10.0 (6.9-13.1)* |  | *13.2 (6.7-20.1)* |
| Unknown/Unstaged | 50-64 | 682 | 87.9 (84.9-90.3) | 750 | 95.1 (93.0-96.6) |
|  | 65-74 | 479 | 79.7 (75.4-83.4) | 302 | 86.7 (81.5-90.5) |
|  | 75-84 | 377 | 76.1 (70.6-80.8) | 198 | 77.7 (69.9-83.7) |
|  | 85-99 | 361 | 56.0 (49.3-62.1) | 115 | 49.1 (37.4-59.7) |
|  | *Difference* |  | *11.8 (6.3-17.8)* |  | *17.4 (11.1-25.3)* |
